# Aging Signals on Chest Radiographs: Association of Chest Radiograph-Derived Age Acceleration With Future Lung Cancer Incidence

**DOI:** 10.64898/2026.03.30.26349022

**Authors:** Yasuhito Mitsuyama, Shannon L Walston, Hirotaka Takita, Kenichi Saito, Daiju Ueda

**Affiliations:** Department of Diagnostic and Interventional Radiology, Graduate School of Medicine, Osaka Metropolitan University, 1-4-3 Asahi-machi, Abeno-ku, Osaka 545-8585, Japan; Smart Data and Knowledge Services Department, German Research Center for Artificial Intelligence (DFKI GmbH), 67663 Kaiserslautern, Germany; Department of Artificial Intelligence, Graduate School of Medicine, Osaka Metropolitan University, 1-4-3 Asahi-machi, Abeno-ku, Osaka 545-8585, Japan; Center for Health Science Innovation, Osaka Metropolitan University, 1-4-3, Asahi-machi, Abeno-ku, Osaka 545-8585, Japan

**Keywords:** Lung Neoplasms, Radiography, Thoracic, Deep Learning, Aging, Risk Assessment

## Abstract

**Purpose:** To evaluate whether chest radiograph–derived age acceleration is associated with incident lung cancer and whether it improves discrimination beyond established lung cancer risk factors.

**Materials and Methods:** This retrospective analysis used prospectively collected data from the Prostate, Lung, Colorectal, and Ovarian Cancer Screening Trial. Baseline digitized chest radiographs from the initial screening year were analyzed using a previously validated deep learning model that estimates chest radiograph–derived age (Xp-age). Age acceleration (AgeAccel) was defined as the residual of Xp-age after calibration to chronological age using a regression model from the development dataset. A 1-year landmark design excluded participants diagnosed with lung cancer or censored within 1 year of baseline. Associations with incident lung cancer were assessed using multivariable Cox proportional-hazards models adjusted for prespecified demographic and clinical predictors, including smoking variables used in the PLCOm2012 risk prediction model. Discrimination was evaluated using the concordance index and 6-year time-dependent area under the receiver-operating-characteristic curve.

**Results:** The analytic cohort included 23,213 participants (mean age, 62.5 years); 790 developed incident lung cancer after the landmark (mean follow-up, 16.7 years). Higher AgeAccel was associated with increased lung cancer incidence (hazard ratio, 1.10 per 1-SD increase; 95% confidence interval: 1.03–1.17); however, addition of AgeAccel to an established risk factor model resulted in minimal change in discrimination (C-index, 0.840 vs. 0.839; time-dependent AUC at 6 years, 0.852 vs. 0.852). Attribution maps emphasized the aortic arch/mediastinal region with similar spatial patterns across smoking and lung cancer strata.

**Conclusion:** Chest radiograph–derived age acceleration was independently associated with future lung cancer incidence.

**Summary:** Chest radiograph–derived age acceleration was associated with future lung cancer incidence in the PLCO cohort after adjustment for established demographic and smoking-related risk factors.

**Key points:**

- In 23,213 participants with baseline chest radiographs and a 1-year landmark design, higher chest radiograph–derived age acceleration was independently associated with incident lung cancer over a mean follow-up of 16.7 years (790 events); the adjusted hazard ratio was 1.10 per 1 standard deviation increase (95% confidence interval, 1.03–1.17; p = 0.003).
- The association between chest radiograph–derived age acceleration and incident lung cancer persisted after full adjustment for smoking status, cigarette consumption, duration, and years since cessation, suggesting that this imaging-based aging phenotype captures a dimension of lung cancer susceptibility beyond smoking exposure and may reflect interindividual variation in thoracic aging that conventional risk factors do not fully represent.

## Introduction

Lung cancer remains a major cause of cancer morbidity and mortality worldwide. In global estimates for 2022, lung cancer accounted for a large share of both incident cancers and cancer deaths (1). Screening with low-dose computed tomography (LDCT) reduces lung-cancer mortality, as shown in the National Lung Screening Trial and the NELSON trial (2,3). Current recommendations endorse LDCT screening for adults at increased risk on the basis of age and smoking exposure, but the population-wise impact of screening depends on identifying those most likely to benefit while minimizing unnecessary downstream evaluation and potential harms (4,5).

Biological aging may help explain why lung cancer develops in some individuals with similar conventional risk profiles and not in others. Aging biomarkers estimate biological age, and age acceleration is commonly defined as the residual after calibration to chronological age (6,7). Using this residual-based approach, Levine and colleagues reported that intrinsic epigenetic age acceleration in blood was associated with subsequent lung cancer in a prospective cohort (6). However, results have varied across cohorts and clock definitions; for example, a nested case–control study within the CLUE II cohort did not support a positive association between several epigenetic age-acceleration measures and lung cancer risk (7). In contrast, other prospective analyses have reported positive associations for some methylation-based aging measures, particularly for lung cancer (8,9). These mixed findings suggest that the relevance of age acceleration to lung-cancer susceptibility may depend on the biologic domain being measured and motivate evaluation of aging signals that are closer to the organ at risk.

The heterogeneity of findings across epigenetic clock studies may in part reflect the organ-distal nature of blood-based biomarkers: DNA methylation measured in peripheral blood cells does not directly capture aging-related structural and inflammatory changes within the thoracic microenvironment most proximate to lung carcinogenesis (10,11). Chest radiography, by contrast, directly images the lung parenchyma, mediastinum, and aortic arch—structures that undergo progressive morphological remodeling with age and that have been independently linked to cancer susceptibility through vascular and inflammatory mechanisms (12,13). Prior work has demonstrated that deep learning models can estimate biological age from chest radiographs, and that the discrepancy between this radiograph-derived age and chronological age correlates with all-cause mortality, cardiovascular outcomes, and chronic disease burden, establishing radiographic appearance as a multidimensional sensor of organismal health (14–17). In parallel, deep learning (DL) applied to chest radiographs can identify smokers at elevated lung cancer risk and predict long-term cancer incidence when combined with clinical data, demonstrating that routine radiographs encode latent risk information that extends beyond conventional imaging interpretation (18–20). Together, these lines of evidence motivate the hypothesis that a chest radiograph–derived aging phenotype—proximal to the organ at risk—may capture a dimension of lung cancer susceptibility not fully represented by smoking exposure, demographic characteristics, or blood-based aging biomarkers.

Whether a chest radiograph–derived aging phenotype—operationalized as age acceleration—is independently associated with incident lung cancer after rigorous adjustment for established clinical and smoking-related risk factors has not been directly tested in a large prospective cohort. Clinical risk models such as PLCOm2012 improve patient screening by integrating demographic characteristics, smoking intensity and duration, years since quitting, and comorbidities, yet they may not capture aging-related susceptibility reflected in imaging (21,22). We therefore evaluated, in the Prostate, Lung, Colorectal, and Ovarian (PLCO) Cancer Screening Trial cohort (19,23), the association between chest radiograph–derived age acceleration (AgeAccel)—defined as the calibrated residual of Xp-age (chest radiograph–derived predicted age, a deep learning–estimated biological age derived from the radiographic appearance) on chronological age—and incident lung cancer using a landmark design to reduce reverse causation and multivariable Cox models adjusted for established lung-cancer risk factors, including predictors used in PLCOm2012 (17,21).

## Materials and Methods

### Study design and data source

The primary objective was to evaluate the association between chest radiograph–derived age acceleration (AgeAccel) and incident lung cancer and to assess whether AgeAccel improves discrimination beyond established predictors. This retrospective analysis used prospectively collected data from the Prostate, Lung, Colorectal, and Ovarian (PLCO) Cancer Screening Trial (19,23), a multicenter randomized trial that evaluated chest radiographic screening for lung cancer. We used a previously published DL approach for Xp-age estimation without fine-tuning or transfer learning (17). This study followed the Declaration of Helsinki. The institutional review board of Osaka Metropolitan University approved the study (Approval Number: 2022-151) and waived additional informed consent because this was a secondary analysis of de-identified PLCO data. We report according to TRIPOD+AI where applicable (24).

### Participants and image acquisition

We used PLCO participants with available baseline chest radiographs and linked clinical and outcome data. In PLCO, the lung-screening examination included standardized chest radiography performed at participating centers according to the trial protocol. We restricted the baseline assessment to digitized screening radiographs obtained in the initial screening year of the PLCO lung-screening protocol. To reduce reverse causation from preclinical lung cancer at baseline, we applied a 1-year landmark design and excluded participants with lung cancer diagnosis or censoring within 1 year after the baseline chest radiography; follow-up for time-to-event analyses began at the landmark. Finally, for the multivariable Cox analysis, we excluded participants with missing data for one or more prespecified covariates (complete-case analysis).

### Outcome definition and follow-up

The primary outcome was incident lung cancer diagnosed after the 1-year landmark. For time-to-event analyses, time zero was defined as 1 year after the baseline chest radiograph. Event time was calculated as the interval from this 1-year landmark to the date of lung cancer diagnosis. Participants who died without lung cancer or remained alive without lung cancer were censored at death or at the end of follow-up, whichever occurred first. Thus, the Cox model estimated the association between baseline AgeAccel and subsequent lung cancer incidence among participants who were alive, uncensored, and lung cancer–free at the 1-year landmark.

### Chest radiograph–derived predicted age (Xp-age) and age acceleration (AgeAccel)

The Xp-age model, detailed in a previous study (17), calculated Xp-age from chest radiographs without requiring additional variables like chronological age or comorbidities. The model was developed and externally validated using 67,099 radiographs from healthy participants. Chest radiographs for that study were downscaled to 320 pixels on the longer side, preserving the aspect ratio, and zero-padded to 320×320. Preprocessing for the current study was performed identically to the original Xp-age study.

To derive an age-acceleration metric, we used a linear regression model based on the Xp-age development data to estimate the expected Xp-age as a function of chronological age. AgeAccel was defined as the residual (observed Xp-age minus expected Xp-age for a given chronological age), such that higher values reflect greater radiographic age acceleration (AgeAccel = Xp-age - [α + β×chronological age]). The regression coefficients α and β were estimated by ordinary least squares in the development dataset using the training and validation splits only (the development test split was not used), consistent with external-application practice.

### Covariates

Covariates were selected a priori based on established lung cancer risk factors and PLCO availability, including age, sex, race, educational level, body-mass index, smoking status (current, former, never), cigarettes per day, smoking duration, years since quitting, chronic obstructive pulmonary disease, personal history of cancer, and family history of lung cancer.

### Model Attribution

Saliency maps were generated for each radiograph using Shapley Additive exPlanations (SHAP) (25). SHAP values were computed with a masking-based approach applied to fixed-size image patches. We summarized model attribution using mean absolute SHAP values and created pixel-wise averaged maps for (A) the overall analytic sample, (B) prespecified baseline age groups (<60 years, 60 to 64 years, 65 to 69 years, and ≥70 years), (C) incident lung cancer status during follow-up (incident lung cancer vs no incident lung cancer), and (D) baseline smoking status (never, former, or current smoker). Two board-certified radiologists reviewed the averaged SHAP maps and recorded qualitative interpretations of hot regions and cold regions, with reference to patterns reported in prior work on chest radiography as a biomarker of aging. The two radiologists reviewed the averaged maps independently and resolved discrepancies by consensus.

### Statistical analysis

We summarized baseline characteristics overall and according to a median split of AgeAccel. We estimated lung cancer–free survival with Kaplan–Meier methods and compared groups with a log-rank test. The proportional-hazards assumption was assessed using Schoenfeld residuals. We fitted two multivariable Cox proportional-hazards models with time zero defined at the 1-year landmark: (1) a base model including prespecified established demographic, clinical, and smoking-related predictors, and (2) an extended model that additionally included AgeAccel. Chronological age and AgeAccel were modeled as continuous variables and scaled to 1-standard-deviation (SD) units; hazard ratios (HRs) are reported per 1-SD increase, with the AgeAccel hazard ratio obtained from the extended model. Participants were censored at death or at the end of follow-up. Discrimination of the base and extended models was evaluated using the concordance index (C-index) and the time-dependent area under the receiver-operating-characteristic curve (AUC) at 6 years after the landmark. The time-dependent AUC at 6 years was defined at 6 years after the 1-year landmark (ie, approximately 7 years after the baseline radiograph) and therefore evaluated discrimination for incident lung cancer occurring within 6 years after the landmark. Confidence intervals for discrimination metrics were estimated by bootstrap resampling. Analyses were conducted in R version 4.4.1. Two-sided p values are reported, with 95% confidence intervals (CI).

## Results

A total of 89,716 radiographs from 25,000 participants were available, from which we restricted the baseline assessment to radiographs obtained in the initial screening year. After applying the 1-year landmark and excluding participants with missing covariates for multivariable adjustment, 23,213 participants remained in the final analytic cohort (Figure 1). During follow-up after the 1-year landmark (mean, 16.7 years; SD, 5.4), 790 participants developed incident lung cancer (Table 1). The mean chronological age at baseline was 62.5 years (SD, 5.4), and the mean Xp-age was 58.8 years (SD, 6.6); the mean AgeAccel was −4.0 years (SD, 4.9) (Table 1). At baseline, 51.0% of participants were men, 87.1% were White, 6.8% had chronic obstructive pulmonary disease, and 11.5% were current smokers (Table 1).

**Figure 1:**
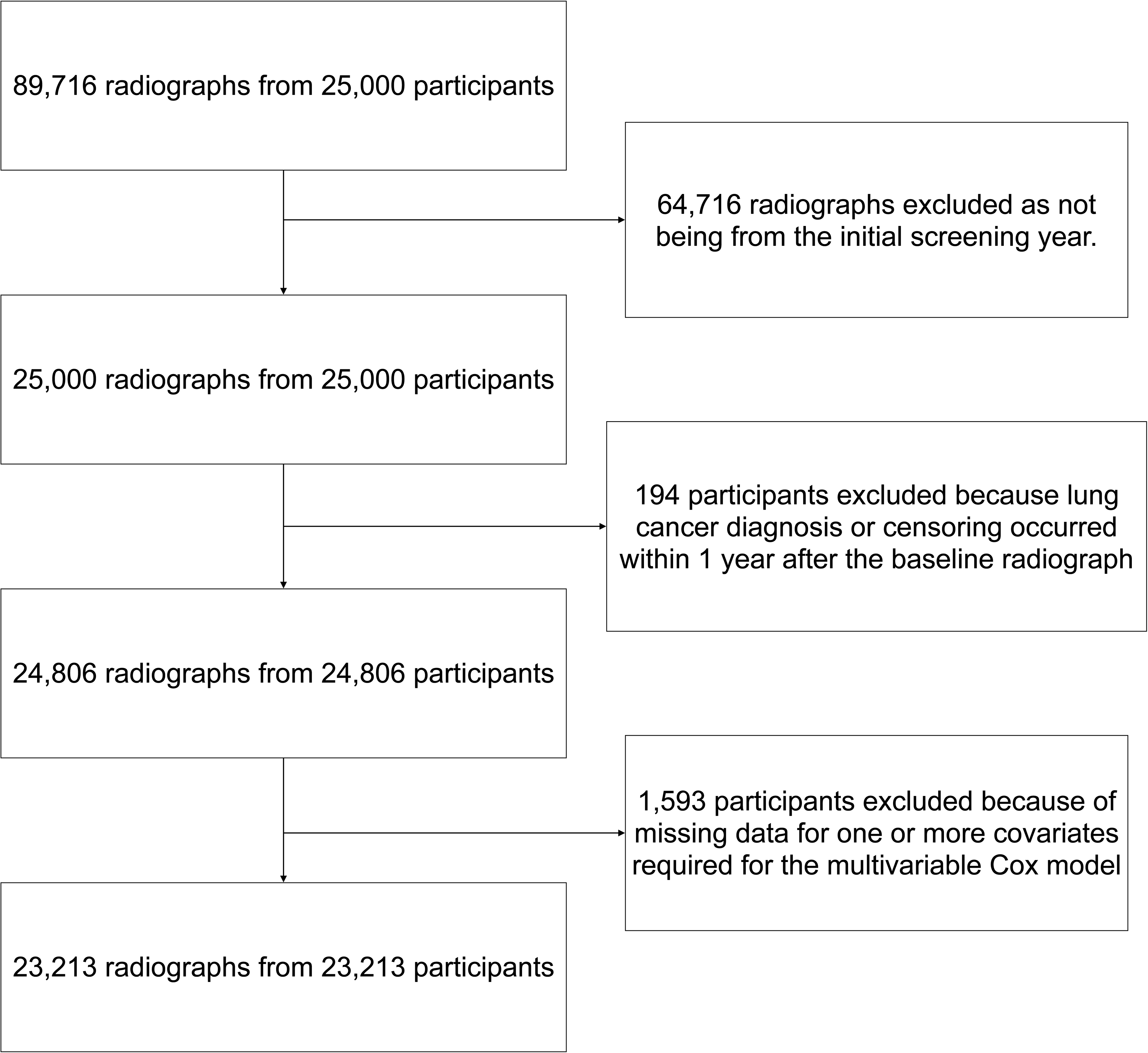
Eligibility flowchart of dataset. This is the selection of participants and chest radiographs from the Prostate, Lung, Colorectal, and Ovarian (PLCO) Cancer Screening Trial. Radiographs not obtained in the initial screening year were excluded. A 1-year landmark design was applied to reduce reverse causation; participants with lung cancer diagnosis or censoring within 1 year after the baseline chest radiography were excluded, and follow-up began at the landmark. Participants with missing data for prespecified covariates required for multivariable adjustment were excluded, yielding the final analytic cohort.

**Table 1:** Baseline Characteristics of the Participants.

|  | Overall | AgeAccel < median | AgeAccel ≥ median |
| --- | --- | --- | --- |
| No. of participants | 23,213 | 11,606 | 11,607 |
| No. of lung cancer events | 790 | 353 | 437 |
| Age, years | 62.5 (5.4) | 63.2 (5.7) | 61.8 (5.0) |
| Xp-age, years | 58.8 (6.6) | 55.6 (6.3) | 62.0 (5.3) |
| AgeAccel, years | -4.0 (4.9) | -7.9 (3.1) | -0.1 (3.0) |
| Follow-up after landmark, years | 16.7 (5.4) | 16.8 (5.3) | 16.6 (5.5) |
| Sex |  |  |  |
| Male | 11,837 (51.0%) | 5,880 (50.7%) | 5,957 (51.3%) |
| Female | 11,376 (49.0%) | 5,726 (49.3%) | 5,650 (48.7%) |
| Race |  |  |  |
| White | 20,228 (87.1%) | 10,283 (88.6%) | 9,945 (85.7%) |
| Black | 1,385 (6.0%) | 655 (5.6%) | 730 (6.3%) |
| Hispanic | 381 (1.6%) | 212 (1.8%) | 169 (1.5%) |
| Asian | 1,031 (4.4%) | 384 (3.3%) | 647 (5.6%) |
| Pacific Islander | 136 (0.6%) | 51 (0.4%) | 85 (0.7%) |
| American Indian | 52 (0.2%) | 21 (0.2%) | 31 (0.3%) |
| Education level |  |  |  |
| 1: Less than high-school graduate | 1,562 (6.7%) | 716 (6.2%) | 846 (7.3%) |
| 2: High school graduate | 5,112 (22.0%) | 2,352 (20.3%) | 2,760 (23.8%) |
| 3: Some training after high school | 2,828 (12.2%) | 1,387 (12.0%) | 1,441 (12.4%) |
| 4: Some college | 5,067 (21.8%) | 2,558 (22.0%) | 2,509 (21.6%) |
| 5: College graduate | 4,074 (17.6%) | 2,138 (18.4%) | 1,936 (16.7%) |
| 6: Postgraduate or professional degree | 4,570 (19.7%) | 2,455 (21.2%) | 2,115 (18.2%) |
| Body-mass index | 27.2 (4.9) | 27.4 (5.1) | 27.0 (4.7) |
| Chronic obstructive pulmonary disease |  |  |  |
| No | 21,639 (93.2%) | 10,929 (94.2%) | 10,710 (92.3%) |
| Yes | 1,574 (6.8%) | 677 (5.8%) | 897 (7.7%) |
| Personal history of cancer |  |  |  |
| No | 22,150 (95.4%) | 11,086 (95.5%) | 11,064 (95.3%) |
| Yes | 1,063 (4.6%) | 520 (4.5%) | 543 (4.7%) |
| Family history of lung cancer |  |  |  |
| No | 20,512 (88.4%) | 10,256 (88.4%) | 10,256 (88.4%) |
| Yes | 2,701 (11.6%) | 1,350 (11.6%) | 1,351 (11.6%) |
| Smoking status |  |  |  |
| Never | 10,202 (43.9%) | 5,463 (47.1%) | 4,739 (40.8%) |
| Former | 10,341 (44.5%) | 5,153 (44.4%) | 5,188 (44.7%) |
| Current | 2,670 (11.5%) | 990 (8.5%) | 1,680 (14.5%) |
| Cigarettes per day | 11.0 (14.2) | 10.3 (14.0) | 11.6 (14.3) |
| Duration of smoking, years | 15.6 (17.3) | 14.1 (16.7) | 17.1 (17.7) |
| Years since quitting, years | 9.0 (12.8) | 9.4 (13.2) | 8.5 (12.4) |

Data are presented as mean (SD) for continuous variables and number (percent) for categorical variables. Follow-up time is shown as years after the 1-year landmark (i.e., time zero for survival analyses was set at 1 year after the baseline chest radiography).

Kaplan–Meier curves showed lower lung cancer–free survival among participants with AgeAccel at or above the median than among those below the median (log-rank p = 0.002) (Figure 2).

**Figure 2:**
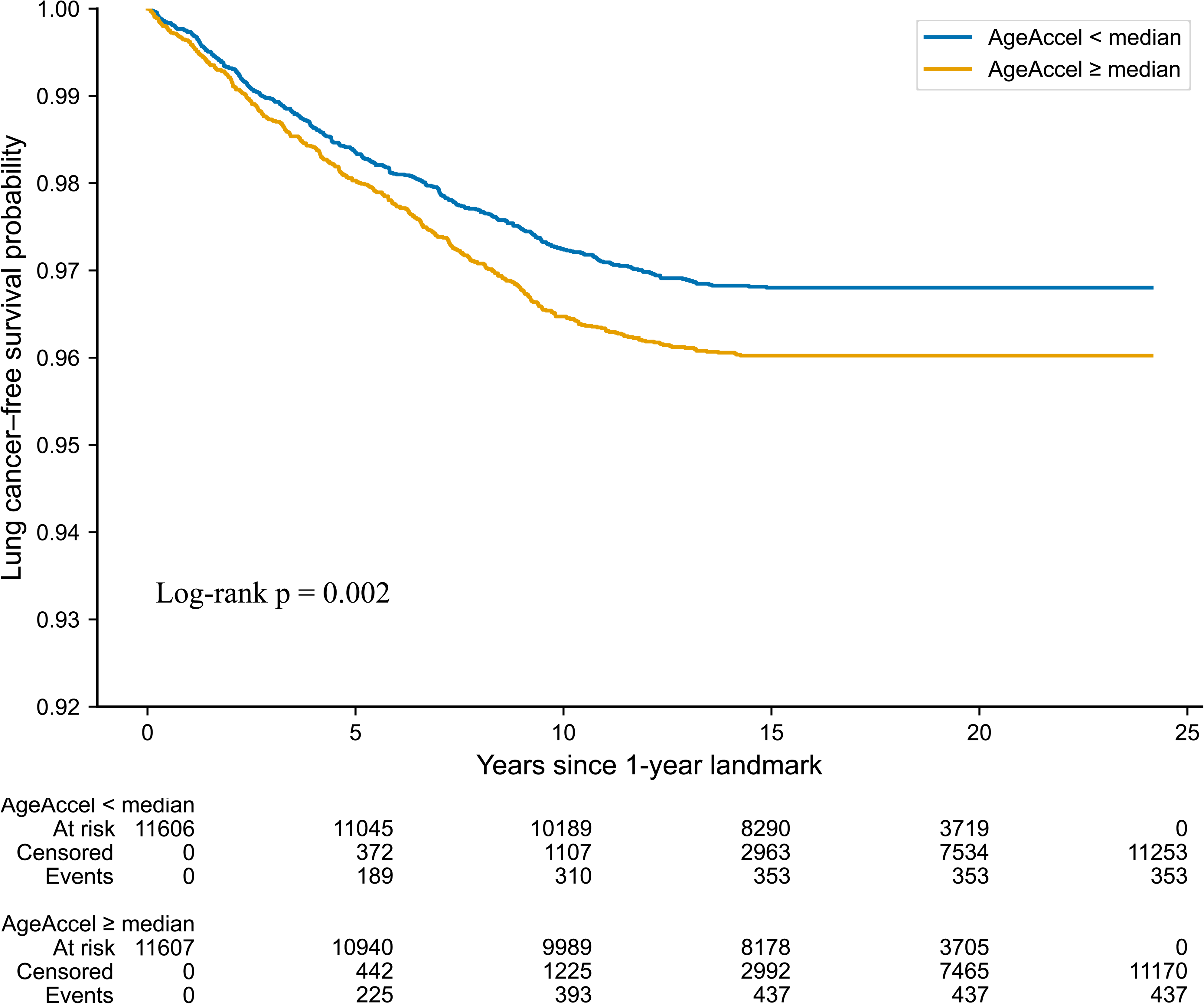
Kaplan-Meier curves and log-rank tests with risk stratification using AgeAccel. Kaplan–Meier curves show lung cancer–free survival from the 1-year landmark after the baseline chest radiography, according to a median split of AgeAccel (AgeAccel ≥ median vs AgeAccel < median). Participants were followed until incident lung cancer or censoring at death or the end of follow-up, whichever occurred first. The p value was calculated with the log-rank test. Numbers at risk are shown below the plot.

In multivariable Cox proportional-hazards models adjusted for prespecified demographic and clinical covariates, higher AgeAccel was associated with a higher risk of incident lung cancer (HR, 1.10 per 1-SD increase; 95% CI, 1.03–1.17) (Table 2, Figure 3). In the same model, chronological age was associated with incident lung cancer (HR, 1.39 per 1-SD increase; 95% CI, 1.30–1.48), as were chronic obstructive pulmonary disease (HR, 1.44; 95% CI, 1.20–1.73) and family history of lung cancer (HR, 1.26; 95% CI, 1.06–1.50) (Table 2, Figure 3). Compared with never smoking, former smoking (HR, 1.25; 95% CI, 1.03–1.52) and current smoking (HR, 2.05; 95% CI, 1.65–2.56) were associated with higher risk, and additional smoking-dose metrics were independently associated with risk (cigarettes per day: HR, 1.02 per cigarette/day increase; 95% CI, 1.01–1.02; smoking duration: HR, 1.03 per year; 95% CI, 1.03–1.04; years since quitting: HR, 0.99 per year; 95% CI, 0.98–1.00) (Table 2).

**Figure 3:**
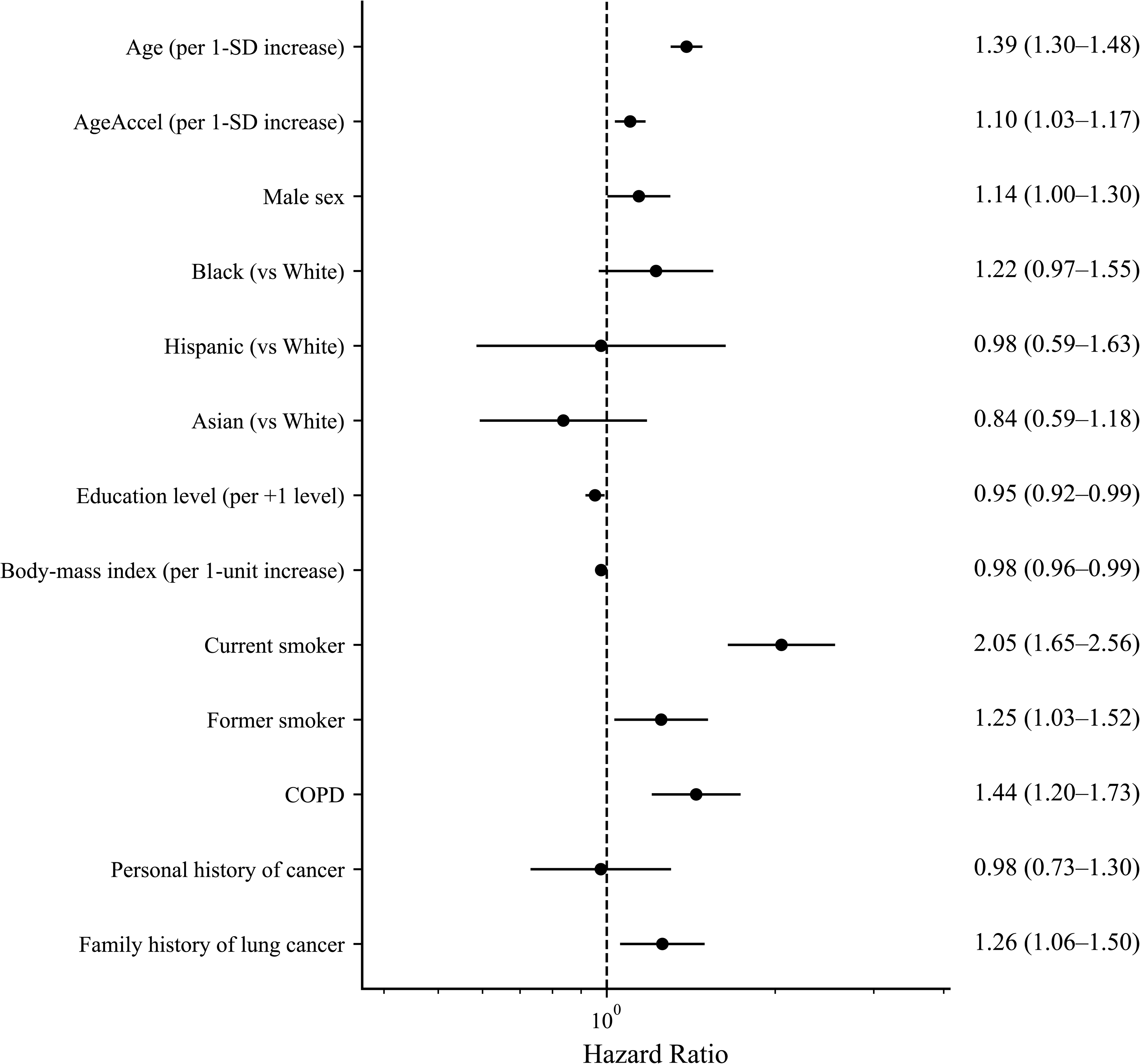
Forest plot of multivariable Cox model hazard ratios. Forest plot showing hazard ratios from the multivariable Cox proportional-hazards model with follow-up starting at the 1-year landmark. Age and AgeAccel are reported per 1-SD increase. Points indicate hazard ratios and horizontal bars indicate 95% confidence intervals. Selected covariates are displayed; full model results are provided in Table 2.

**Table 2:**
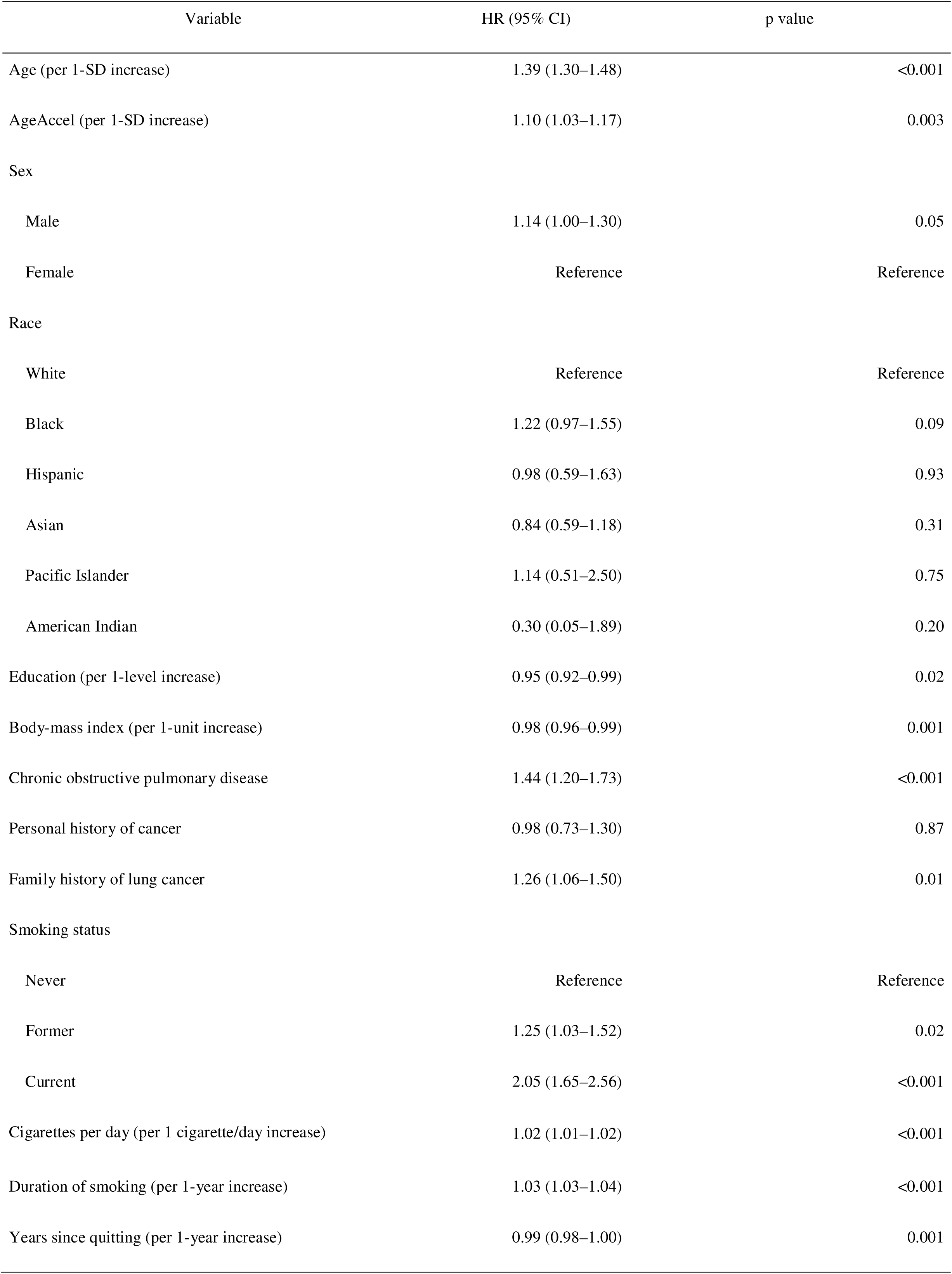
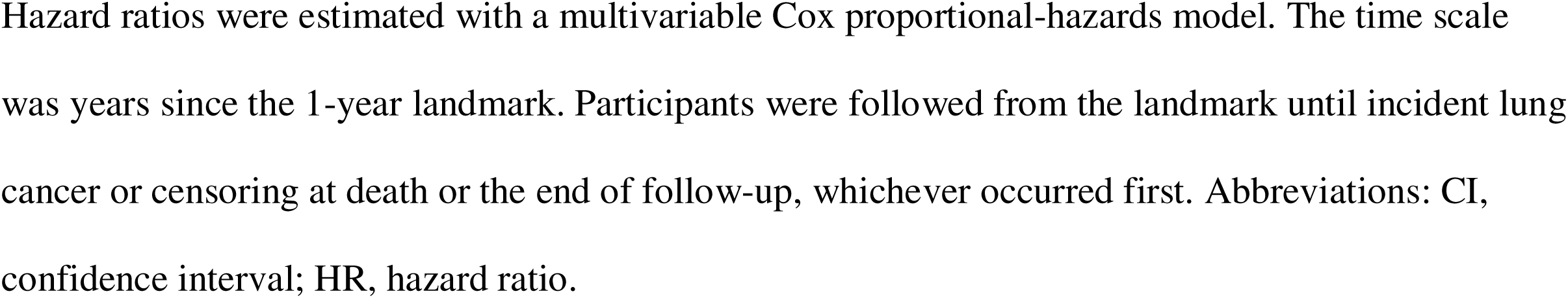
Association Between Chest Radiograph–Derived Age Acceleration and Incident Lung Cancer.

| Variable | HR (95% CI) | p value |
| --- | --- | --- |
| Age (per 1-SD increase) | 1.39 (1.30–1.48) | <0.001 |
| AgeAccel (per 1-SD increase) | 1.10 (1.03–1.17) | 0.003 |
| Sex |  |  |
| Male | 1.14 (1.00–1.30) | 0.05 |
| Female | Reference | Reference |
| Race |  |  |
| White | Reference | Reference |
| Black | 1.22 (0.97–1.55) | 0.09 |
| Hispanic | 0.98 (0.59–1.63) | 0.93 |
| Asian | 0.84 (0.59–1.18) | 0.31 |
| Pacific Islander | 1.14 (0.51–2.50) | 0.75 |
| American Indian | 0.30 (0.05–1.89) | 0.20 |
| Education (per 1-level increase) | 0.95 (0.92–0.99) | 0.02 |
| Body-mass index (per 1-unit increase) | 0.98 (0.96–0.99) | 0.001 |
| Chronic obstructive pulmonary disease | 1.44 (1.20–1.73) | <0.001 |
| Personal history of cancer | 0.98 (0.73–1.30) | 0.87 |
| Family history of lung cancer | 1.26 (1.06–1.50) | 0.01 |
| Smoking status |  |  |
| Never | Reference | Reference |
| Former | 1.25 (1.03–1.52) | 0.02 |
| Current | 2.05 (1.65–2.56) | <0.001 |
| Cigarettes per day (per 1 cigarette/day increase) | 1.02 (1.01–1.02) | <0.001 |
| Duration of smoking (per 1-year increase) | 1.03 (1.03–1.04) | <0.001 |
| Years since quitting (per 1-year increase) | 0.99 (0.98–1.00) | 0.001 |

Hazard ratios were estimated with a multivariable Cox proportional-hazards model. The time scale was years since the 1-year landmark. Participants were followed from the landmark until incident lung cancer or censoring at death or the end of follow-up, whichever occurred first. Abbreviations: CI, confidence interval; HR, hazard ratio.

Discrimination of the base model that included established clinical risk factors was high (C-index, 0.839 [95% CI, 0.826–0.854]; time-dependent AUC at 6 years, 0.852 [95% CI, 0.833–0.871]) and changed minimally with the addition of AgeAccel (C-index, 0.840 [95% CI, 0.827–0.855]; time-dependent AUC at 6 years, 0.852 [95% CI, 0.833–0.872]) (Table 3). Across prespecified age strata, differences in the C-index and 6-year AUC between the base model and the model including AgeAccel were small, and the estimated association between AgeAccel and incident lung cancer was directionally similar but less precisely estimated in older age groups (Table 3). Specifically, the HR per 1-SD increase in AgeAccel was 1.12 (95% CI, 0.99–1.28) for participants younger than 60 years, 1.11 (95% CI, 0.99–1.24) for those 60 to younger than 65 years, 1.10 (95% CI, 0.98–1.24) for those 65 to younger than 70 years, and 1.12 (95% CI, 0.94–1.34) for those 70 years of age or older (Table 3).

**Table 3:** Discrimination of Risk Models With and Without Chest Radiograph–Derived Age.

|  | C-index (95% CI) | Time-dependent AUC (95% CI) | AgeAccel HR (per 1-SD) (95% CI) | p value |
| --- | --- | --- | --- | --- |
| Overall |  |  |  |  |
| Base model | 0.839 (0.826–0.854) | 0.852 (0.833–0.871) | NA | NA |
| The model with AgeAccel | 0.840 (0.827–0.855) | 0.852 (0.833–0.872) | 1.10 (1.03–1.17) | 0.003 |
| Age <60 years |  |  |  |  |
| Base model | 0.855 (0.835–0.890) | 0.859 (0.830–0.897) | NA | NA |
| The model with AgeAccel | 0.857 (0.838–0.893) | 0.860 (0.834–0.897) | 1.12 (0.99–1.28) | 0.07 |
| Age 60-<65 years |  |  |  |  |
| Base model | 0.820 (0.798–0.851) | 0.820 (0.793–0.857) | NA | NA |
| The model with AgeAccel | 0.820 (0.796–0.852) | 0.819 (0.792–0.856) | 1.11 (0.99–1.24) | 0.07 |
| Age 65-<70 years |  |  |  |  |
| Base model | 0.829 (0.810–0.856) | 0.850 (0.826–0.875) | NA | NA |
| The model with AgeAccel | 0.830 (0.811–0.856) | 0.850 (0.827–0.877) | 1.10 (0.98–1.24) | 0.11 |
| Age ≥70 years |  |  |  |  |
| Base model | 0.808 (0.776–0.846) | 0.833 (0.797–0.875) | NA | NA |
| The model with AgeAccel | 0.808 (0.778–0.847) | 0.833 (0.798–0.875) | 1.12 (0.94–1.34) | 0.20 |

The base model included established clinical risk factors; the second model additionally included AgeAccel. Discrimination was summarized with the concordance index (C-index) and time-dependent area under the receiver-operating-characteristic curve (AUC) at 6 years after the landmark. Hazard ratios for AgeAccel are shown per 1-SD increase. Abbreviations: AUC, area under the curve; CI, confidence interval; HR, hazard ratio; NA, not applicable.

In averaged SHAP maps, two radiologists identified a hot region near the aortic arch and a cold region in the lower lung fields in the overall sample, and these patterns varied across age strata, with the aortic-arch hot region becoming more prominent in older age groups and the lower-lung cold region becoming more prominent in younger age groups (Figure 4). In contrast, when maps were stratified by incident lung cancer status and by smoking status, the spatial distribution of hot and cold regions appeared qualitatively similar across strata, with no marked differences in the overall pattern (Figure 4).

**Figure 4:**
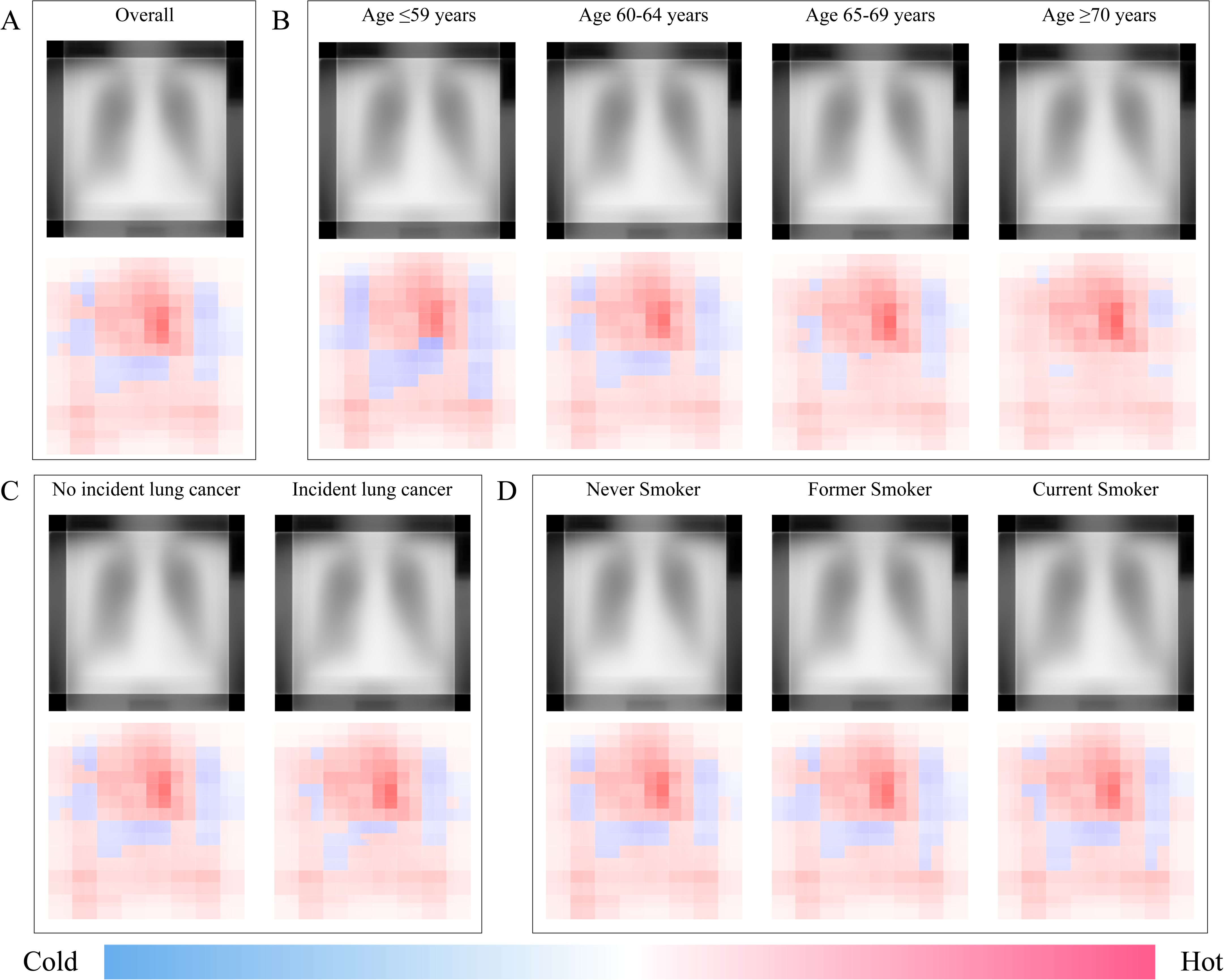
Saliency maps derived from Shapley Additive exPlanations. (A) The overall averaged saliency maps. (B) Averaged saliency maps stratified by chronological age groups (≤59, 60–64, 65–69, and ≥70 years). (C) Averaged saliency maps stratified by incident lung cancer status. (D) Averaged saliency maps stratified by smoking status.

## Discussion

Our study showed that chest radiograph–derived age acceleration was associated with incident lung cancer in the PLCO cohort, independent of established clinical and smoking-related risk factors. In the 1-year landmark analysis, higher AgeAccel was independently associated with increased lung cancer risk (HR, 1.10 per 1-SD increase), with corresponding differences in lung cancer–free survival by AgeAccel median split (log-rank p = 0.002). The C-index and time-dependent AUC at 6 years changed minimally with the addition of AgeAccel. Accordingly, rather than immediate gains in patient prognostication, AgeAccel serves as an indicator of physiologic changes in the thorax, including mediastinal and vascular structures, visible on chest radiographs.

The novelty of our study lies in operationalizing a chest radiograph–based aging phenotype as an “age-acceleration” biomarker and testing its association with lung cancer incidence using an epidemiologic framework that explicitly controls for confounding by established risk factors and mitigates reverse causation. Prior work on lung-cancer risk from chest radiographs has focused largely on direct prediction of cancer risk or screening eligibility, demonstrating that DL models can identify smokers at high risk even when chest radiography is not effective as a screening test for mortality reduction (18). Separate works on radiographic age estimation have shown that an AI-derived chest radiograph age (CXR-Age) predicts long-term mortality and that Xp-age and its discrepancy from chronological age correlate with chronic disease burden, supporting the use of radiography as a potential “aging sensor” in routine care (14–17). By combining these lines of inquiry, our study reframes information in the chest radiograph as a quantitative aging phenotype and evaluates whether that phenotype is linked to incident lung cancer over long follow-up within PLCO.

Our findings also support a clinically meaningful implication: lung cancer risk may reflect not only smoking exposure and demographic risk factors but also measurable age-related physiologic and morphologic changes that can be captured on a widely available imaging test. Risk prediction models such as PLCOm2012 integrate age, smoking duration and intensity, years since quitting, and comorbidities such as COPD and have been shown to improve efficiency of screening selection compared with categorical eligibility thresholds (21,22). Nonetheless, these models may not fully capture heterogeneity in lung cancer susceptibility—differences in pulmonary reserve, systemic inflammation, and cumulative injury—that may influence carcinogenesis (26–28). Aging biomarkers derived from blood DNA methylation have been associated with cancer risk in some studies, including lung cancer, though results have not been uniform (6,7,29). Against this background, a chest radiograph–derived aging phenotype may offer complementary information that is proximal to the organ at risk and available at scale. Although the incremental change in discrimination in our analysis was small, the consistent direction of association after adjustment suggests that radiographic aging may capture a dimension of vulnerability that is not fully accounted for by conventional predictors, and it may be informative for understanding why individuals with similar smoking histories experience different cancer risks.

AgeAccel may reflect thoracic aging features—particularly vascular and mediastinal remodeling and age-related changes in lung appearance—that may correlate with lung cancer susceptibility. Prior work on chest radiograph–based age phenotypes, including CXR-Age and Xp-age, has suggested that age-related signals are prominent around mediastinal contours and the aortic arch region and that lower lung fields may show relative negative attribution in averaged maps, consistent with cardiopulmonary remodeling across the lifespan(14,17). In our averaged SHAP maps, two board-certified radiologists identified a hot region near the aortic arch and a cold region in the lower lung fields in the overall sample, and the intensity of these regions varied across age strata in a pattern similar to that reported previously, with a more prominent aortic-arch hot region in older participants and more prominent lower-lung cold regions in younger participants (17). Notably, the spatial distribution of hot and cold regions appeared qualitatively similar when stratified by incident lung cancer status and by smoking status, suggesting that the model attribution patterns primarily reflect a generalized radiographic aging phenotype rather than radiographic features specific to lung cancer or smoking exposure. This interpretation is compatible with the possibility that radiographic age acceleration captures systemic or vascular aging—processes that share pathways with carcinogenesis—rather than acting as a surrogate for occult cancer on the baseline chest radiograph (12,13).

Several limitations should be considered. First, although we used a 1-year landmark design to reduce reverse causation from occult cancers present at baseline, preclinical disease could still have influenced radiographic appearance beyond 1 year; additional sensitivity analyses with longer lag periods would further strengthen causal interpretation. Second, residual confounding remains possible, particularly from measurement error in smoking variables and from unmeasured occupational or environmental exposures. Third, our analyses were conducted as complete-case analyses for multivariable models; if missingness is informative, effect estimates and performance metrics may be biased, and multiple imputation could be considered. In addition, discrimination metrics were evaluated in the same cohort used for model fitting and may therefore be optimistic; future work should include stronger internal validation and external evaluation. Fourth, generalizability may be limited by imaging acquisition conditions and population characteristics in PLCO, and external validation in more contemporary and diverse cohorts would be important, particularly if the intended use is screening triage. Finally, age-stratified analyses are inherently less precise, and interpretation should be cautious in light of multiplicity and reduced sample size in some strata; these analyses are best presented as assessments of consistency rather than as claims of differential effect.

In conclusion, chest radiograph–derived age acceleration was independently associated with incident lung cancer in PLCO, suggesting that radiographic aging phenotypes may capture clinically relevant vulnerability beyond conventional risk factors. These results extend prior work showing that chest radiographs can encode latent risk information and that radiographic age estimates correlate with long-term outcomes and disease burden (14,17,18). The immediate incremental gain in discrimination from adding AgeAccel to a traditional risk model was small, indicating that near-term clinical utility may lie more in mechanistic and phenotypic insight than in stand-alone risk prediction. Future studies should prioritize external validation, sensitivity analyses that further address reverse causation and competing risks, and careful evaluation of potential clinical workflows—such as augmenting individualized risk models for LDCT screening or identifying subsets for targeted prevention—while adhering to contemporary reporting standards for prediction modeling and medical imaging AI.

## Data Availability

All data produced in the present study are available upon reasonable request to the authors.

## Acknowledgements

This study used data (and images) from the Prostate, Lung, Colorectal, and Ovarian (PLCO) Cancer Screening Trial, accessed through the National Cancer Institute (NCI) Cancer Data Access System (CDAS) under approved project number PLCOI-1285. We thank the NCI for making the PLCO data available to the research community, and we acknowledge the PLCO screening center investigators and staff, as well as the staff of Information Management Services, Inc. and Westat, Inc. Most importantly, we thank the PLCO trial participants for their invaluable contributions. The statements contained herein are solely those of the authors and do not necessarily represent, or imply concurrence or endorsement by, the National Cancer Institute.

This work was supported by Japan Science and Technology Agency (JST) as part of Adopting Sustainable Partnerships for Innovative Research Ecosystem (ASPIRE), Japan, Grant Number JPMJAPJST2403, JST BOOST, Japan, Grant Number JPMJBS2401, and JSPS KAKENHI, Japan, Grant Number JP24K18804.

